# Patient-derived ovarian cancer organoids mimic clinical response and exhibit heterogeneous inter- and intrapatient drug responses

**DOI:** 10.1101/2019.12.12.19014712

**Authors:** Chris J. de Witte, Jose Espejo Valle-Inclan, Nizar Hami, Kadi Lõhmussaar, Oded Kopper, Celien P.H. Vreuls, Trudy N. Jonges, Paul van Diest, Luan Nguyen, Hans Clevers, Wigard P. Kloosterman, Edwin Cuppen, Hugo J.G. Snippert, Ronald P. Zweemer, Petronella O. Witteveen, Ellen Stelloo

## Abstract

**Purpose:** There remains an unmet need for preclinical models to enable personalized therapy for ovarian cancer (OC) patients. Recently, patient-derived organoid (PDO) cultures of patients with OC have been established that faithfully represent the histopathological features and genomic landscape of the patient tumor. In this study, we evaluate the capacity of OC PDOs to predict clinical drug response and functional consequences of tumor heterogeneity.

**Experimental design:** 36 genomically characterized PDOs from 23 patients with known clinical histories were exposed to chemotherapeutics and targeted drugs.

**Results:** OC PDOs maintained genomic features of the original tumor lesion and recapitulated patient response to neoadjuvant carboplatin and paclitaxel combination treatment, according to distinct clinical outcomes (histopathological, biochemical and radiological). PDOs displayed inter-as well as intrapatient drug response heterogeneity, which could in part be explained by genetic aberrations. All PDOs were resistant to PARP-inhibitors, in accordance with homologous recombination pathway fidelity and genome-wide mutation context. *KRAS, BRAF* and *NRAS* mutation status predicted response to BRAF-inhibitor vemurafenib and pan-HER-inhibitor afatinib, and explained differential response among four PDOs derived from distinct tumor locations of an individual patient. Importantly, PDO drug screening identified sensitivity to at least one drug for the majority of patients (88%).

**Conclusions:** OC PDOs are a valuable preclinical model system that can provide insights in drug response for individual patients with OC, complementary to genetic testing. Generating PDOs of multiple tumor locations can improve clinical decision making and increase our knowledge on genetic and drug response heterogeneity.

## Introduction

Epithelial ovarian cancer (OC) is characterized by the development of chemotherapy resistance and poor survival. Overall survival for patients with OC has only slightly improved over the past decades, despite developments in the field such as optimized surgical tumor resection, administration of (hyperthermic) intraperitoneal chemotherapy and introduction of targeted treatments such as PARP-inhibitors (1). While most patients with OC respond well to initial treatment, the majority will develop recurrent disease within the first two years and become resistant to chemotherapy. In the setting of recurrent disease, a wide range of chemotherapeutic and targeted drugs is available. PARP-inhibitors are indicated for patients who experienced complete or partial response to previous platinum treatment, irrespective of *BRCA1/2-*mutation status (2–4). Still, *BRCA1/2*-mutation carriers experience more benefit from PARP-inhibition compared to patients with homologous recombination (HR)-proficient tumors (5). However, for the majority of relapsed patients and drugs, no genetic markers are available to predict response. These patients might benefit from patient-derived model systems that can be employed to test response to drugs prior to treatment in the clinic.

Traditionally, OC drug response has been studied in 2D-cell lines and xenografts. 2D-cell lines offer a relatively cheap and quick model system, suitable for high-throughput drug screening; while patient-derived xenografts offer the potential to study tumor drug response in a living organism, but are not suitable for high-throughput drug screening experiments (6). In the past decade patient-derived organoids (PDOs) have been established, a 3D-cell culture model system that maintains cellular heterogeneity of healthy tissues and tumors. Recently, PDOs of OC were established which represent the genomic features of the original tumors (7–9). Furthermore, a drug screening comparison between 2D-cultures and PDOs of OC revealed that cytostatic drug efficacy was dependent on the employed culture system; PDO drug responses correlated better with genomic aberrations compared to 2D-cell cultures (9). To employ the organoid system to guide treatment choice in the clinic, it is vital that the correlation between PDO drug response and clinical drug response is established. To this extent, prospective clinical trials have been performed with PDOs of patients with colorectal cancer, in which *in vitro* drug screening recapitulated patient response to chemotherapy and targeted drugs (10,11). For OC, we and others previously provided anecdotal evidence on the correlation between clinical and PDO drug response (7,8,12), but direct comparisons are still limited.

When predicting treatment response, genetic heterogeneity should be considered. Epithelial OC, especially the high-grade serous subtype, is a heterogeneous disease with widespread inter- and intrapatient genetic heterogeneity (13,14). A virtue of the PDO model system is the possibility to study genetic and phenotypic tumor heterogeneity (15).

In this study, we systematically assessed if *in vitro* drug response of OC PDOs correlates to patients’ clinical response to chemotherapeutics. We studied inter- and intrapatient drug response heterogeneity to a wide range of chemotherapeutics as well as targeted drugs, and linked differential drug response to genetic variation.

## Materials and methods

### Patient inclusion, sample and clinical data collection

For this study we included patients with epithelial OC. Each patient signed informed consent and was able to withdraw her consent at any time. Tumor samples, ascites and blood samples were gathered between January 2016 and September 2019 at the University Medical Center Utrecht, and Leiden University Medical Center, The Netherlands. Patient data and tissue collection was performed according to the guidelines of the European Network of Research Ethics Committees (EUREC) following European, national and local law. The Institutional Review Board of the UMC Utrecht (IRB UMCU) approved the biobanking protocol: 14-472 HUB-OVI. Clinical data was collected from the patient files. Patient samples were derived at primary disease during primary debulking surgery or interval debulking surgery, or adnex extirpation procedures. Upon recurrence, tissue was collected during (laparoscopic) surgery performed for treatment or diagnostic purposes, or ascites was collected during palliative drainage procedures. For the comparison of PDO drug response and clinical response, solely patients that underwent interval debulking surgery were considered.

### Organoid derivation and culture

Organoids were derived from tumor samples of patients with OC and cultured according to our previously described protocol (7). PDO names are informative of histological subtype, patient and tumor location. Histological subtype: HGS/LGS=high/low-grade serous adenocarcinoma, HG=high-grade adenocarcinoma, SBT/MBT=serous/mucinous borderline tumor, MC=mucinous adenocarcinoma, CCC=clear cell carcinoma, END=endometrioid carcinoma. The first number indicates the patient, the second number indicates tumor location.

### *In vitro* PDO drug response testing and data-analysis

PDO drug response testing was performed as previously described (7). In short, PDOs were exposed to drugs in varying concentrations and to controls (DMSO, ABT-263/navitoclax) for 120 hours in 384-well plates, after which ATP levels were measured with the Cell-Titer Glo2.0 assay (Promega BV). All screens were performed in technical replicates. Biological replicates were performed in a subset of PDOs and drugs (supplementary table 6) to investigate biological variation. Results were normalized to vehicle (DMSO = 100%) and baseline control (ABT-263/navitoclax 20 μM). Data were analyzed using GraphPad Prism 6. Drug dose-response curves were visualized using linear regression analysis (setting: log(inhibitor) vs. normalized response --variable slope; least squares (ordinary) fit; no constraints). Concentrations where 50% cell viability (IC50-value) was reached were interpolated. The area under the curve (AUC) was approximated between the lowest and highest concentrations screened in the actual assay with the trapezoid rule for numerical integration.

### Therapeutic agents

Selleckchem: alpelisib (BYL719) catalog no. S2814; adavosertib (MK-1775) catalog no. S1525; afatinib (BIBW2992) catalog no. S1011; AZD8055 catalog no. S1555; carboplatin catalog no. S1215; gemcitabine catalog no. S1714; MK-2206 catalog no. S1078; niraparib (MK-4827) catalog no. S2741; olaparib (AZD2281, Ku-0059436) catalog no. S1060; paclitaxel catalog no. S1150; pictilisib (GDC-0941) catalog no. S1065; Rucaparib (AG-014699,PF-01367338) catalog no. S1098; Vemurafenib (PLX4032, RG7204) catalog no. S1267. Bio-connect: flavopiridol (146426-40-6) catalog no. HY-10005. Active Biochem: cobimetinib (GDC-0973) catalog no. A-1180.

### Clinical drug response measures

Histopathological response was assessed with the chemotherapy response score (CRS) after three cycles of neoadjuvant chemotherapy, according to the guidelines described by Bohm et al (16). All available hematoxylin and eosin stained slides of each tumor location from which we established PDOs were assessed for tumor purity. The slide with the most tumor per location was subsequently blinded scored by two certified pathologists (PvD, CV), as CRS-1 (no or minimal pathological response), CRS-2 (appreciable pathological response) or CRS-3 (complete or near-complete pathological response). In case of disagreement consensus was reached. Radiological response was assessed according to the RECIST criteria for solid tumors (version 1.1) (17). A score for each patient was obtained, from best to worst response: complete response (CR), partial response (PR), stable disease (SD), or progressive disease (PD). Biochemical response was measured by assessing response and timing of normalization (<35kU/L) of biomarker cancer antigen 125 (CA-125) (18). For progression-free survival (PFS) a cut-off of six months was employed; since patients with less than six months PFS are predicted to be resistant to subsequent platinum-treatment. For overall survival 17 months was taken as a cut-off, based on survival data of a recent cohort of patients (2015-2016) with HGSOC stage IIIC and IV disease by the Dutch Cancer Registration. Seventeen months after diagnosis, 50% of patients with HGSOC stage IV were still alive (supplementary table 3).

### DNA isolation and whole genome sequencing (WGS)

DNA was isolated with the DNeasy Qiagen kit (PDOs and blood samples) and Genomic Tip Qiagen kit (tumor samples), supplemented with RNase treatment. Fresh frozen tumor material obtained through biopsy procedures was processed with the QiaSymphony DSP DNA kit for low input. For DNA library preparation, 500–1,000 ng of DNA was used. Subsequently, whole-genome paired-end sequencing (2x 150 bp) was performed on Illumina HiSeq X Ten and NovaSeq 6000 to a median coverage of 31X (range 24-45X).

### WGS data analysis

WGS data were processed using the Hartwig Medical Foundation (HMF) somatic mutation workflow. We installed the pipeline (v4.8) locally using GNU Guix with the recipe from https://github.com/UMCUGenetics/guix-additions. Full details and pipeline description are explained in detail by Priestley et al. (19)(https://github.com/hartwigmedical/pipeline). Briefly, sequence reads were mapped against human reference genome GRCh37 with Burrows-Wheeler Alignment (BWA-MEM) (v0.7.5a) (20). Indel realignment and base recalibration was performed with the Genome Analysis Toolkit (GATK, v3.8.1) (21). Somatic single nucleotide variants (SNVs) and small insertions and deletions were called with Strelka (v1.0.14) (22). The functional effect of the somatic SNVs and indels were predicted with SnpEff (v.4.3) (23). Somatic structural variants (SVs) were called with GRIDSS (v1.8.0) (24).

Copy number alterations (CNAs) were called with PURPLE (25). PURPLE also assesses tumor purity. In case of low tumor purity, a quality flag was raised (“NO_TUMOR”; supplementary table 9). For tumor samples MC-3.2, MC-3.3, MBT-2.1 and MC-1.2, based on manual verification, a wrong ploidy level was automatically selected by PURPLE. Therefore, and due to the impossibility of manually correcting the ploidy selection in PURPLE, we ran Control-FREEC (v. 11.0) (26) instead on all samples (tumor and PDO) from those patients. Telomeric and centromeric regions were masked for visualization.

For samples CCC-1, END-1.1, END-1.2, HGS-22 and HGS-23 no normal reference sample was available for somatic mutation calling. In these cases, germline SNV calling was performed with GATK (21) and only SNVs with a “HIGH” or “MODERATE” effect as predicted by SnpEff were considered. Similarly, germline SV calling was performed using GRIDSS and SV calls were filtered against the SV Panel of Normals from the HMF analysis pipeline, which can be found in https://resources.hartwigmedicalfoundation.nl. Since PURPLE requires tumor-normal pairs, CNA calling for these five samples was performed individually with Control-FREEC (v. 11.0) (26).

### Assessment of homologous recombination (HR) status

To identify HR-deficient samples, *BRCA1* and *BRCA2* as well as other genes in the HR-pathway (*BARD1, BRIP1, CHEK2, FANCA, PALB2, RAD50, RAD51(B/C/D)*) were assessed for biallelic inactivation, incorporating both germline and somatic WGS data. Biallelic inactivation was defined as: a deep deletion (i.e. full loss of both alleles); or Loss-Of-Heterozygosity (LOH) in combination with (i) a known pathogenic/likely pathogenic variant according to ClinVar (https://www.ncbi.nlm.nih.gov/clinvar/; GRCh37, database date 2018-12-07), or (ii) a frameshift, nonsense or essential splice variant as annotated by SnpEff (http://snpeff.sourceforge.net/; v4.1h). Additionally, CHORD (**C**lassifier of **HO**mologous **R**ecombination **D**eficiency; Nguyen et al, in preparation) was employed to classify PDO samples as HR-proficient or -deficient based on the presence of genome-wide somatic mutation contexts (primarily deletions with flanking microhomology and 1-100kb structural duplications). Samples without a germline reference sample were excluded from CHORD evaluation.

### Selection of genes with known drug-gene interactions

The Drug Gene Interaction database (DGIdb) was utilized as a resource to obtain a list of genes that have a known interaction with drug response (27). WGS data of PDOs were checked for SNVs, SVs and CNAs in DGIdb genes, in case differential drug response was observed within related PDOs. Homo-polymer regions were excluded. To identify significantly amplified and deleted genes we applied stringent criteria adopted from Priestley et al (19). An amplification was defined as [minimum exonic copy number > three times the sample ploidy], while a deletion was defined as [minimum exonic copy number <0.5 times the sample ploidy]. Related samples were regarded genetically heterogeneous on copy number level, if they presented with different copy-number states (amplified vs neutral vs deleted). Furthermore, differential response among related samples was only considered if the ploidy-corrected copy number levels were >10% apart.

### Statistical analysis

Descriptive statistics including mean, SD and SEM were conducted with R (software package, version 3.5.0). The significance level for 95% confidence interval was set to α=0.05. The Pearson correlation test was applied to evaluate the correlation between replicate experiments. The Wilcoxon signed-rank test was applied for the comparison of clinical response groups. The means of two technical replicates of each sample at all measured drug concentrations were compared between clinical response groups (CRS-1 vs −2, RECIST SD vs PR, no CA-125 normalization vs normalization, PFS <6 months vs >=6 months, OS <17 months vs >=17 months). We corrected for multiple testing with the Bonferroni method (alpha = 0.05 / 5 (tests)), resulting in a statistically significant threshold of p=0.01.

### PDO availability

Available PDOs are cataloged at www.hub4organoids.eu and can be requested at. Distribution of PDOs to third parties will have to be authorized by the IRB UMCU at the request of the HUB to ensure compliance with the Dutch ‘medical research involving human subjects’ act.

### Data availability

BAM files of WGS data are made available through controlled access at the European Genome-phenome Archive (EGA) which is hosted at the EBI and the CRG (https://ega-archive.org), under dataset accession number EGA: EGAD00001005707. Data access requests will be evaluated by the UMCU Department of Genetics Data Access Board (EGAC00001000432) and transferred on completion of a material transfer agreement and authorization by the IRB UMCU at the request of the HUB to ensure compliance with the Dutch ‘medical research involving human subjects’ act.

## Results

### PDOs can be (rapidly) established from different OC subtypes

In total, we included 36 PDOs (of which 29 were established previously (7)), derived from 23 patients with different histological subtypes of OC who underwent primary or interval debulking surgery or ascites drainage (supplementary table 1). PDO sample names are informative of histological subtype as well as patient (first number) and tumor location (second number).The majority of PDOs in our biobank were thoroughly characterized and biobanked prior to drug testing, to establish a reliable platform. However, in order to incorporate PDO-based drug response prediction in clinical care, PDO establishment and screening must be executed within a short time span. As a pilot experiment, we successfully established and rapidly screened organoids from a patient with recurrent disease (HG-26; supplementary figure 1). Within 20 days of tumor collection the response to six therapies became available.

### PDOs retained genomic features of the original tumor lesion

First, we compared the genomic profiles of PDOs and the tumors they were derived from. An average of 8,290 and 10,358 SNVs were identified in the parental tumor specimen and their matched PDO, respectively. On average 67% of variants were shared between the tumor and PDO, 6% of the SNVs were unique to the tumor, and 27% to the PDO (supplementary figure 2). Assessment of CNAs demonstrated comparable copy-number states in the majority of pairs (supplementary figure 3). HGS-3.1, LGS-2.2 and MC-2.1 presented with a much higher number of SNVs than their matched tumor specimen and considerable CNA dissimilarities within PDO-tumor pairs. These exceptions are likely due to a high degree of normal cell contamination in the tumor samples, which was confirmed by PURPLE, a purity ploidy estimator (supplementary table 9) (25). In general, PDOs are enriched for tumor cells, whereas tumor samples are heterogeneous, representing a mix of tumor cells and normal cells. For tumor samples with sufficient tumor content, PDOs retained the genomic features of the original tumor lesions.

### PDO drug response correlates to patients’ clinical response

Next, we evaluated the potential of PDOs to reflect patients’ drug response to chemotherapy. For this we selected all PDOs that were derived at interval debulking surgery from patients with HGS OC, with known clinical histories (supplementary table 1 (clinical comparison) and 2). Seven PDOs (derived from five patients) were exposed to carboplatin and paclitaxel combination treatment *in vitro* and we could directly compare their response to the patient’s clinical response. Related samples HGS-3.1 and HGS-3.2 were most sensitive to carboplatin and paclitaxel combination treatment (AUC=0.37 and 0.29), while HGS-24 was resistant (AUC=0.88) (figure 1). These PDO drug responses showed a statistically significant correlation (p<0.01) with clinical response as measured by histopathological (chemotherapy response score (CRS)), biochemical (normalization of serum biomarker CA-125) and radiological (RECIST) responses (figure 1; supplementary figure 4). PDOs derived from tumor locations with no or minimal histopathological response (CRS=1) were less sensitive to carboplatin and paclitaxel combination treatment compared to PDOs derived from tumor locations with appreciable pathological response (CRS=2) (p=5.821e-05, Wilcoxon signed-rank test) (16). Biochemically, clinical drug response is measured according to the response criteria and timing of normalization of CA-125 (17,18). Even though all patients exhibited CA-125 response during primary treatment, PDOs derived from patients who did not reach CA-125 normalization during primary treatment were less sensitive to the chemotherapeutics compared to PDOs from the patient in whom CA-125 levels normalized (<35kU/L, p=0.0004). Radiological response was assessed according to the RECIST criteria (version 1.1) (17), comparing imaging data at initiation of chemotherapy to imaging data prior to interval debulking based on CT-scanning. PDOs derived from patients with RECIST stable disease were less sensitive to carboplatin and paclitaxel combination treatment compared to PDOs from patients with RECIST partial response (p=0.0092). To compare long-term clinical response to PDO response, recurrence and survival were assessed. All patients experienced recurrent disease within four to 14 months after the last primary treatment, and 6-month progression-free survival (PFS) did not correlate to PDO drug response. After 17 months 50% of patients with FIGO stage IV HGS OC are still alive (supplementary table 3); only one out of five patients (HGS-24) in our cohort lived shorter than 17 months, and this PDO exhibited strongest resistance to carboplatin and paclitaxel combination treatment.

**Figure 1.**
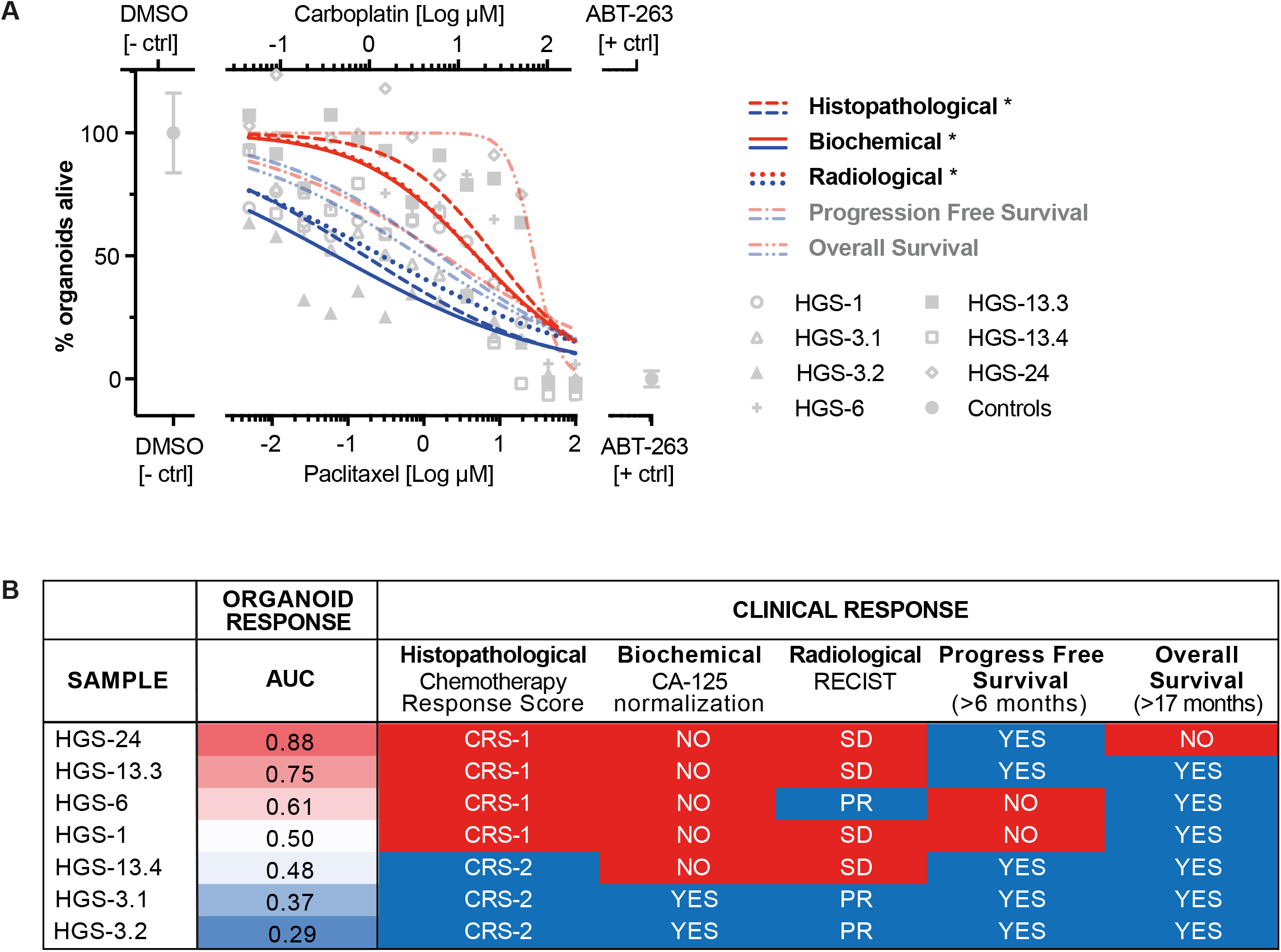
OC PDO drug response correlates with clinical drug response. A) Drug dose-response curves of OC PDOs for carboplatin and paclitaxel combination treatment dichotomized into clinical response groups. Dose response curves are normalized to positive (navitoclax, ABT-263) and negative controls (DMSO). Upper x-axis: carboplatin drug concentrations, lower x-axis: paclitaxel drug concentrations. Non-linear regression analysis: log(inhibitor) vs. normalized response -- Variable slope fit. Red=clinically resistant, blue=clinically sensitive. Histopathological tumor response: CRS1=no or minimal response vs CRS2=appreciable response; p=5.821e-05. Biochemical response: no normalization (<35 kU/L) of serum CA-125 during primary treatment vs normalization; p=0.0004. Radiological response: stable disease (SD) vs partial response (PR) according to RECIST criteria; p=0.0092. *Statistically significant difference between sensitive and resistant group according to Wilcoxon signed-rank test corrected for multiple testing (p<0.01). B) Overview of PDO drug response (area under the curve (AUC)) versus all clinical response measures, ordered from most resistant to most sensitive based on AUC values.

### PDOs exhibit interpatient drug response heterogeneity which correlates partially with their genetic makeup

Next, we investigated the response of all PDOs (N=36) to a broader range of drugs and drug combinations (3-19 per PDO, on average 13; supplementary table 5), including chemotherapeutics and targeted drugs. Drugs were selected based on clinical practice or evaluation in clinical trials for ovarian cancer or solid tumors in general. Drug response experiments were performed in technical replicates and replicate AUC values highly correlated (R^2^=0.87) (supplementary figure 5). PDOs were considered resistant if the drug concentration that reduced viability of >50% of cells (IC50-value) was higher than the concentration achievable in patient plasma (concentration steady state, maximum concentration (C*ss*/C*max*; supplementary table 4) (28,29).

Divergent responses were observed to chemotherapeutic drugs carboplatin (platinum/alkylating agent), paclitaxel (taxane/antimicrotubule agent) and gemcitabine (pyrimidine antagonist) (figure 2A-C; supplementary table 5). A minority of PDOs was sensitive to carboplatin (7/31, 23%) and paclitaxel monotherapy (5/31, 16%), while sensitivity to gemcitabine was observed in most PDOs (29/35, 83%). Response also correlated with OC histological subtype, all LGS-samples showed resistance to paclitaxel and carboplatin monotherapy (except for response to carboplatin in LGS-3.1), while a sensitive response was restricted to HG(S)-samples. For certain PDOs, combination treatment with two chemotherapeutic drugs resulted in lower IC50 values compared to treatment with the single drugs (supplementary table 5). Our results, for example, showed that carboplatin and paclitaxel treatment alone had minimal effect on LGS-3.1 with an IC50-value of 1.46 and >2.5 log μM, respectively, while the IC50-value of carboplatin and paclitaxel was reduced to 0.56 and −0.34 log μM in the combination treatment.

**Figure 2.**
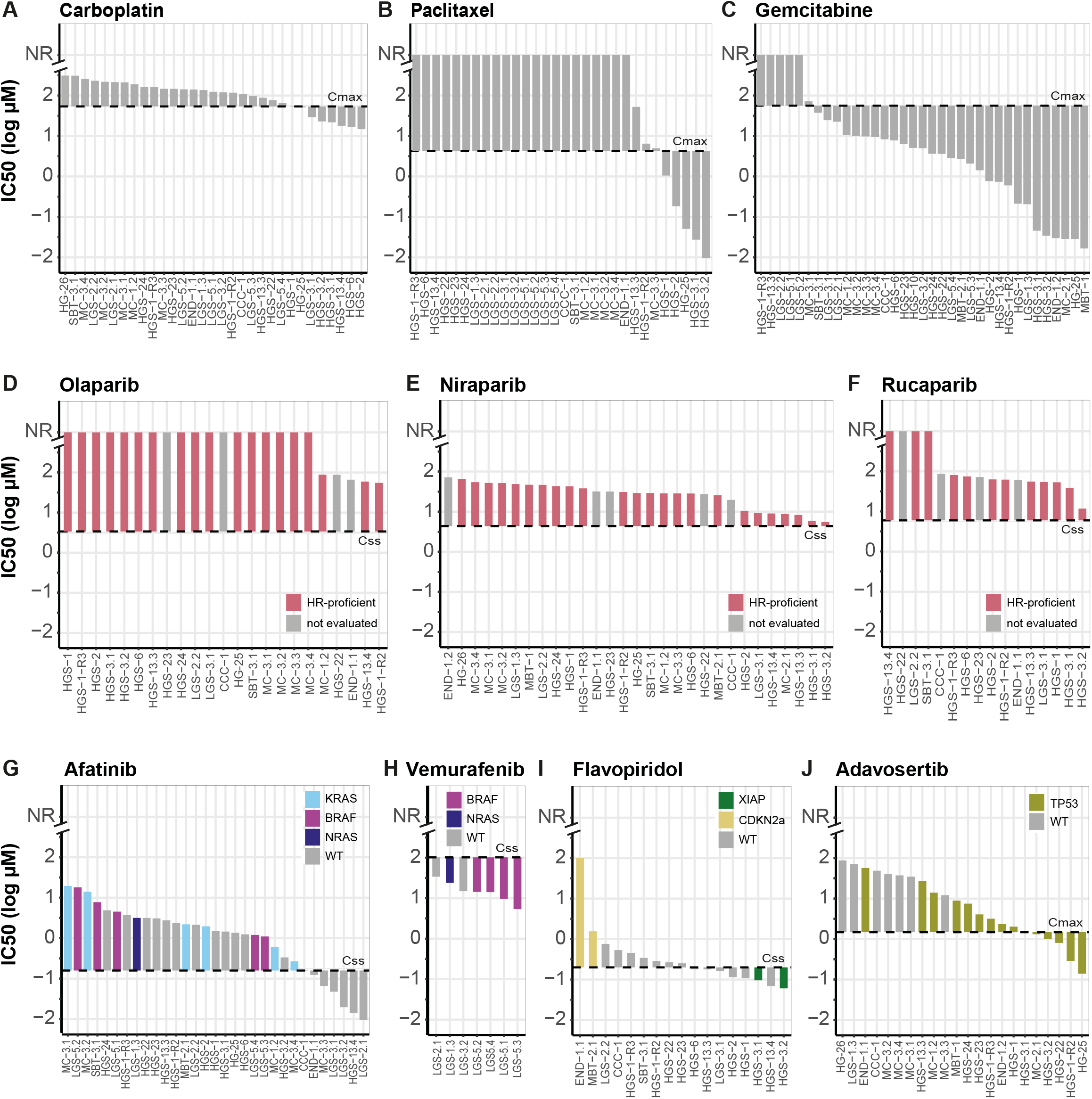
OC PDOs exhibit interpatient heterogeneity in response to chemotherapy and targeted drugs. Waterfall plots with IC50-values (extracted from dose-response curves) of OC PDOs for chemotherapeutics and targeted drugs. The steady state (C*ss*) or maximum (C*max*) in vivo plasma concentrations are indicated with the dotted line (supplementary table 3). Bars north of the dotted line represent resistant samples, bars south of the dotted line represent sensitive samples. A-C. Response to chemotherapeutics carboplatin (A), paclitaxel (B) and gemcitabine (C). D-F. Response to PARP-inhibitors olaparib (D), niraparib (E) and rucaparib (F). All PDOs were resistant to the PARP-inhibitors which correlated with their HR-proficient genetic make-up (no biallelic hit in HR-related genes). Not evaluated = no CHORD evaluation due to missing normal reference. G-J. Response to targeted drugs afatinib (G), vemurafenib (H), flavopiridol (I) and adavosertib (J) could partly be explained by genetic aberrations (color-coded) (supplementary table 7). WT=wildtype for the genes mentioned in each panel. NR=IC50-value not reached.

The responses to targeted drugs revealed differences and similarities between PDOs which in part correlated to their genetic makeup. For example, all PDOs were resistant to the PARP-inhibitors olaparib, niraparib, and rucaparib (figure 2D-F), consistent with the absence of biallelic inactivation of *BRCA1* and *BRCA2*, and other genes involved in homologous recombination (e.g. *CHEK2, FANCA, PALB2, RAD50, RAD51(B/C/D);* supplementary table 8 and 9). Additionally, HR-classifier CHORD (Nguyen et al, in preparation) classified all samples as HR-proficient based on genome-wide somatic mutation contexts (supplementary figure 6). As expected, *BRAF*-, *KRAS*-, and *NRAS*-mutant PDOs showed resistance to pan-HER-inhibitor afatinib (12/25; figure 2G) and sensitivity to BRAF-inhibitor vemurafenib (5/7; figure 2H). Alterations in *CDKN2A* and *XIAP*, known to affect response to CDK-inhibitor flavopiridol, were present in our cohort (30–32). *CDKN2A* was affected in the two PDOs most resistant to flavopiridol: MBT-2.1 showed loss of both alleles and END-1.1 harbored a nonsense variant (p.(R58*); figure 2I), while copy number loss of *XIAP* was observed in two of the flavopiridol-sensitive PDOs (figure 2I). Sensitivity to WEE1-inhibitor adavosertib correlated with *TP53* mutation status. All *TP53* wildtype PDOs (N=7) showed resistance, while *TP53* mutants (N=15) were distributed among both the resistant and sensitive PDOs (figure 2J). For the remaining drugs, alpelisib, AZD-8055, MK-2206, and pictilisib, no known genotype and drug response phenotype correlations were observed.

Subsequently, we evaluated for each individual patient how many of the tested monotherapies (3-13 per patient) remained as potential treatments based on an IC50-value smaller than the achievable concentration in patient plasma (C*ss*/C*max*). In case of multiple tumor locations per patient, all test results were considered. A predicted sensitive response to at least one (and maximum five) drug(s) was observed for 88% of patients, except for HGS-1-R3, MC-3 and HG-26 in which all of the 13, seven and three tested monotherapies yielded a resistant response in at least one of their PDOs (figure 3; supplementary table 5).

**Figure 3.**
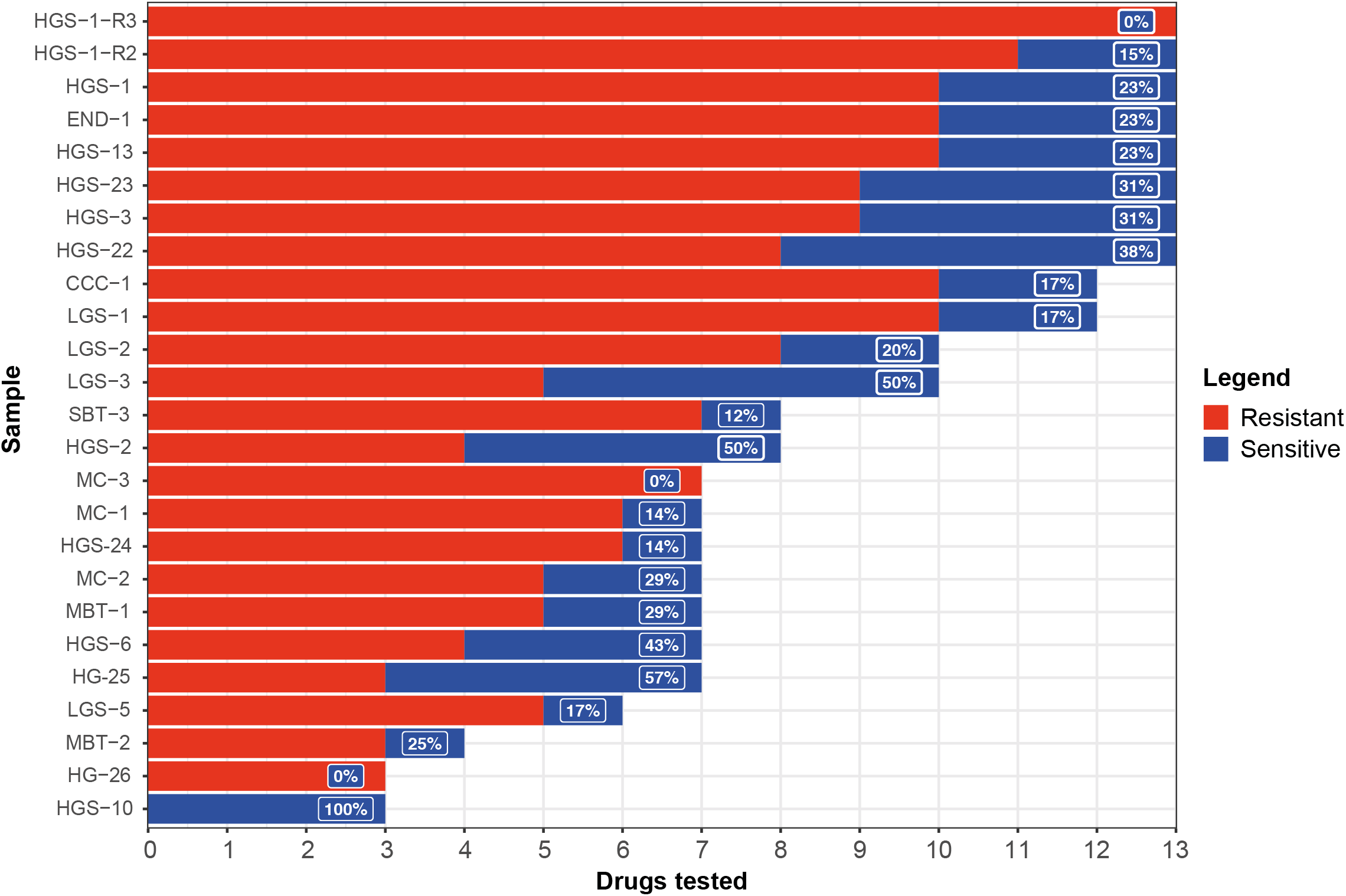
Overview of OC PDO response to single drugs per patient. Overview of the number of monotherapies tested per patient (3-13), classified as predicted sensitive or resistant, based on the IC50-value relative to the in vivo plasma concentration (C*ss*/C*max*, supplementary table 4, 5). For the majority of patients (88%) sensitivity to at least one tested drug was identified. For patients with organoids derived from multiple tumor locations, results from al tested samples were considered. Red=resistant, blue=sensitive.

### PDOs derived from individual patients revealed intrapatient drug response heterogeneity

In addition to assessing interpatient drug response heterogeneity, we examined intrapatient drug response heterogeneity. For seven individual patients, two to four PDOs were derived from distinct cancer lesions at a single time point. For one additional patient, three PDOs were derived at subsequent time points (supplementary table 1, heterogeneity comparison). To set a threshold for differential drug response, we first assessed the extent of biological variability. We observed low drug response variability across biological replicates (N=84) with an IC50-value correlation coefficient of R^2^=0.82 and mean IC50-fold change of 2.5±1.5 (range=1.0-7.3) (supplementary figure 7; supplementary table 6), therefore, a ten-fold change in IC50-value was chosen as a stringent cut-off for differential drug response.

While homogeneous responses were observed to a subset of drugs and drug combinations; carboplatin combined with gemcitabine, adavosertib, or olaparib, carboplatin, olaparib, niraparib, rucaparib, alpelisib, AZD-8055, flavopiridol, pictilisib, and vemurafenib (supplementary figure 8), all related PDOs exhibited differential drug response to at least one drug, as defined by a >10-fold change in IC50-value (figure 4). In the seven patients of whom multiple PDOs were derived at the same time point, differential response to mono-treatment was observed 11/36 times (31%). Importantly, in six cases, one of the samples yielded a sensitive response whereas a related sample yielded a resistant response.

**Figure 4.**
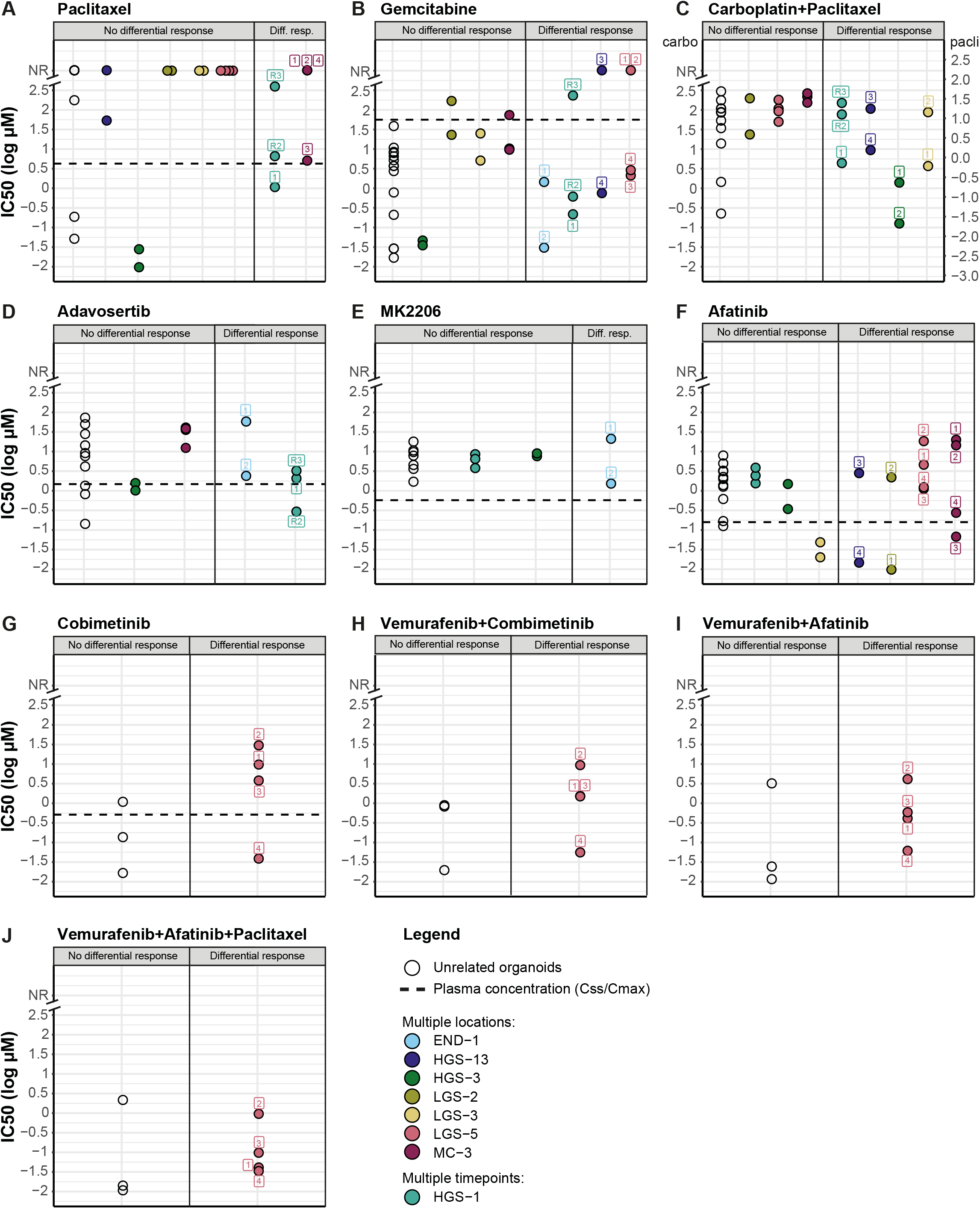
OC PDOs exhibit intrapatient heterogeneity in response to chemotherapy and targeted drugs. IC50-values (extracted from dose-response curves) for drugs that elicit a differential drug response in at least one patient with multiple OC PDOs: paclitaxel (A), gemcitabine (B), carboplatin+paclitaxel (C), adavosertib (D), MK2206 (E), afatinib (F), cobimetinib (G), vemurafenib+cobimetinib (H), vemurafenib+afatinib (I), vemurafenib+afatinib+paclitaxel (J). Differential drug response is defined as >10 fold-change in IC50-value within related samples. Left panel: unrelated and related samples without differential response. Right panel: related samples that exhibited differential response. A color code for each patient is shown. The dotted line indicates the steady state (C*ss*) or maximum (C*max*) in vivo plasma concentrations for all single drug treatments (supplementary table 4).

To examine the impact of intratumor genetic heterogeneity on phenotypic heterogeneity, we assessed genetic variants in genes that are known or predicted to interact with drugs according to the drug-gene interaction database resource (DGIdb; supplementary table 7) (30). HGS-13.3, LGS-2.2, LGS-5.2, MC-3.1 and MC-3.2 PDOs were markedly less sensitive to the pan-HER-inhibitor afatinib compared to their related PDOs (figure 4F). Despite meeting the criteria of differential response, all four *BRAF*-mutant LGS-5 PDOs were classified as resistant to afatinib with an IC50-value above the steady state concentration of −0.8 log μM. The remaining related PDOs with differential response (HGS-13, LGS-2, MC-3) were classified as both sensitive and resistant to afatinib. We observed a direct genotype and drug response phenotype correlation for the four PDOs derived from a patient with a mucinous OC (MC-3). The two most resistant PDOs (MC-3.1 and MC-3.2) harbored a *KRAS* hotspot mutation (p.G12V), whereas the other resistant PDO MC-3.4 harbored two different *KRAS* mutations (p.L19F and p.Q61E, both reported to have an attenuated phenotype compared to hotspot mutations (33,34) and the sensitive PDO MC-3.3 was *KRAS* wildtype (WT) (supplementary table 8). *KRAS* mutations were independently confirmed with Sanger sequencing (supplementary figure 9).

LGS-5 PDOs also exhibited different sensitivities to gemcitabine and the MEK-inhibitor cobimetinib (figure 4B, 4G). LGS-5.1 and LGS-5.2 were both resistant to gemcitabine, whereas LGS-5.3 and LGS-5.4 were sensitive. LGS-5.4 was also sensitive to cobimetinib, while the other LGS-5 PDOs were resistant. All LGS-5 PDOs were largely genetically identical, and no variants or copy number changes were identified that explained differential response to these drugs (supplementary table 8, 9).

HGS-13.3 PDOs revealed a >10-fold higher IC50-value compared to HGS-13.4 PDOs for gemcitabine, the combined carboplatin and paclitaxel treatment and afatinib (figure 4B, 4C, 4F, supplementary table 5). Consistent with previous findings on the effect of copper-efflux pumps on chemotherapy sensitivity (35–37), copy number losses of *ATP7A* and *ATP7B* were identified in the HGS-13.4 (supplementary table 9). Additionally, six other genes previously associated with chemotherapy response (*EIF4EBP1, EDNRB, NAT2, TLE3, BRCA2* and *NRG1*) (38–40), exhibited different copy-number states between HGS-13.3 and −13.4 which may also have contributed to the observed differential response to gemcitabine and combined carboplatin and paclitaxel treatment (supplementary table 9).

END-1 PDOs, both derived from distinct parts of the tumor lesion in the same ovary, demonstrated differential drug response to gemcitabine, WEE1-inhibitor adavosertib and AKT-inhibitor MK-2206 (figure 4B, 4D, 4E). We identified genetic alterations in *WWOX, ERBB2* and *HRAS* that might have contributed to the observed differential response (supplementary table 8, 9) (41–45). However, even though END-1.2 achieved the lowest IC50-values for all three drugs, both END-1.1 and END-1.2 were classified as sensitive to gemcitabine and resistant to MK-2206 and adavosertib (figure 4B, 4D, 4E).

HGS-3.1 and LGS-3.1 displayed drug responses that were very similar to their related PDOs (figure 4; supplementary table 5). In these related PDOs, differential response was only observed for the combined carboplatin and paclitaxel treatment, while drug responses were similar to carboplatin and paclitaxel mono-treatment. Two carboplatin-response associated genes (30), *CLCN6* and *MTHFR*, exhibited copy number loss in the sensitive PDO LGS-3.1 (supplementary table 9). Functional studies have not focused on *CLCN6* and chemotherapy response, but have shown additive effects of *MTHFR*-inhibition and chemotherapeutic drugs (46).

Additionally, drug response heterogeneity was examined in a patient from whom PDOs were obtained at multiple time points. HGS-1 was derived from primary chemosensitive disease and HGS-1-R2 and HGS-1-R3 were derived from recurrent chemoresistant disease, and together these PDOs reflected the clinical course of the patient. HGS-1-R2/R3 were less sensitive to the mono- and combination treatment of carboplatin and paclitaxel compared to HGS-1 (figure 4A, 4C; supplementary table 5). Although HGS-1-R2 and HGS-1-R3 were derived from ascites collected within a timeframe of one month, differential sensitivity was observed to paclitaxel, gemcitabine and adavosertib (figure 4A, 4B, 4D).

Moreover, we assessed if SVs (including gene fusions) in genes from the DGIdb resource could be linked to differential drug response. In the related PDOs derived from the eight patients that exhibited differential drug response, no SVs were identified that could explain phenotypic heterogeneity. In addition to genetic heterogeneity in genes reported to influence drug response by the DGIdb, related PDOs also exhibited varying degrees of genome-wide heterogeneity at both the SNV and CNA levels (supplementary figure 10). The average number of unique SNVs and CNAs in all related PDOs were 24% (2 to 70%) and 17% (0 to 100%), respectively. Considerable genomic heterogeneity at SNV level was observed in LGS-2 and MC-3 (30-70% unique SNVs per PDO), while these PDOs exhibited differential drug response to only one/two drugs. On the other hand, END-1 and LGS-5 had the lowest degree of genomic heterogeneity (2-14% unique SNVs per PDO) and exhibited heterogeneous response to three drugs. In conclusion, we did not observe a direct correlation between genome-wide heterogeneity and differential drug response.

## Discussion

We have performed drug screening on 36 PDOs derived from 23 patients comprising all major OC histopathological subtypes. OC PDOs resembled the tumors they were derived from, with an average overlap of 67% of SNVs and similar CNA profiles. PDOs generated at interval debulking recapitulated clinical response to first-line carboplatin and paclitaxel combination treatment for histopathological (p=5.821e-05), biochemical (p=0.0004) and radiological (p=0.0092) outcomes.

Diverse responses to registered drugs for OC were observed among PDOs. Limited sensitivity to first-line carboplatin (7/31, 23%) and paclitaxel (5/31, 16%) treatment was observed, compared to sensitivity in the majority of patients to second-line gemcitabine treatment (29/35, 83%). All PDOs exposed to PARP-inhibition were found to be resistant, in line with HR-proficiency classification based on WGS data. Response to targeted drugs under clinical investigation could partly be explained by genetic variation; resistance to afatinib (12/25, 48%), resistance to adavosertib (7/17, 41%), response to flavopiridol (N=4/17, 24%) and sensitivity to vemurafenib (5/7, 71%). Importantly, we identified a sensitive response to at least one tested drug in nearly all patients (22/25, 88%). Since not all PDOs were exposed to the same number of drugs (3-13 monotherapies tested per patient), this is likely an underrepresentation. Finally, intrapatient tumor heterogeneity assessment in seven patients with organoids derived from multiple tumor locations, revealed differential response to at least one drug for all patients, indicating the importance to evaluate multiple tumor locations.

In a systematic approach, we showed that PDO drug response correlated with several clinical response measures. This included histopathological assessment of tumor regression according to a three-tier method (CRS) (16), which is recommended for assessment of response to neoadjuvant therapy (47). Histopathological grading of tumor regression offers the advantage to study each tumor site separately, as opposed to patient-wide measures of response such as CA-125 and RECIST. While Bohm *et al*. (16) previously reported that the prognostic significance of the CRS on omental tumor lesions was greater than on primary tumor sites, we applied it to all tumor locations where organoids were derived from. In this study, we present a correlation between CRS and PDO drug response to carboplatin and paclitaxel combination treatment.

In order to bring PDO-based drug response assessment to the clinic, PDO establishment and drug screening needs to be performed within a short timeframe, preferably limited to the diagnosis-treatment interval. In line with previous studies (8,48–50), we demonstrated that rapid PDO establishment and drug screening is feasible. To further validate the predictive value of PDOs, we plan to undertake a prospective trial in which organoids will be derived from both primary and recurrent tumors and tested for response to drugs provided in the clinic, while clinical response is systematically monitored.

Considering intrapatient drug response heterogeneity, derivation of organoids from multiple tumor locations of individual patients, may further improve treatment allocation (51). Although sequencing studies have shown that OCs display extensive inter- and intratumor heterogeneity on a genetic level (13,14), we could only partially link inter- and intratumor heterogeneous drug responses to genetic heterogeneity. Additionally, some of the CNAs that we identified in genes reported to be related to drug response by the DGIdb might be non-contributive passenger events, given the high frequency of CNAs in high-grade serous OC. Therefore, follow-up studies with increased sample sizes and deeper sequencing are required to decipher drug response associations with the candidate genes identified here, and to discover novel resistance mechanisms. Importantly however, the lack of complete correlation between genetic and functional testing at this point, stresses the need for functional testing in addition to genetic testing to improve clinical decision making.

The establishment of a larger collection of OC PDOs will provide the opportunity to determine comprehensive, clinically useful genotype-phenotype correlations. When a large collection including drug response data is available, treatment stratification can potentially be performed based on genomic or transcriptomic characteristics of specific PDO subtypes, which could make organoid derivation dispensable in the future (51). This transition requires an accurate classification of drug-sensitive and -resistant PDOs. Similar to previous studies (28,29), OC PDOs were considered sensitive if the drug concentration that reduced viability of >50% of cells was lower than the concentration achievable in patient plasma (C*ss*/C*max*). However, the *Css/Cmax* will vary between patients and is not necessarily the concentration that is achieved in the tumor (52,53). In addition, sometimes patients require dose adjustments due to adverse events which also affect the drug concentration achievable in both plasma and tumor. Therefore, it should be taken into account that tumors predicted to be sensitive based on the PDO drug response may clinically not respond. Prospective clinical trials comparing clinical to PDO drug response, should be complemented with plasma drug level measurements to further elucidate the relation between *in vitro* and clinical sensitivity.

To conclude, OC PDOs provide a valuable preclinical model system to guide treatment choice in the clinic as it satisfies the following criteria; 1) PDOs genetically resemble the original tumor from which they are derived, 2) PDO drug response reflects patients’ clinical response, and 3) PDO establishment and drug screening can be performed within a short timeframe. Generating and testing PDOs of multiple tumor locations will offer insights in differential drug response as a result of tumor heterogeneity. This information could improve treatment stratification and reduce the development of drug resistance. Complementary PDO drug screening and genomic analysis allows the linkage of genotypes with drug sensitivity patterns to identify candidate biomarkers for drug response.

## Data Availability

BAM files of whole-genome sequencing data are made available through controlled access at the European Genome-phenome Archive (EGA) which is hosted at the EBI and the CRG (https://ega-archive.org), under dataset accession number EGA: EGAD00001005707. Data access requests will be evaluated by the UMCU Department of Genetics Data Access Board (EGAC00001000432) and transferred on completion of a material transfer agreement and authorization by the IRB UMCU at the request of the HUB to ensure compliance with the Dutch ‘medical research involving human subjects’ act.

https://ega-archive.org

## Acknowledgements

We thank members of the Kloosterman and Cuppen laboratories for helpful discussions; Anne Snelting of the Utrecht Platform for Organoid Technology (U-PORT; UMC Utrecht) for patient inclusion and tissue acquisition; Maaike Vreeswijk and Lise van Wijk (Leiden University Medical Center) for providing ovarian cancer tissues for PDO culturing; Vera Deneer for input on clinical pharmacokinetics; Utrecht Sequencing Facility and Hartwig Medical Foundation for providing sequencing service and data; Hans Bos for acquiring funding; and the Dutch Cancer Registration (IKNL) for providing survival data. This work was supported by Gieskes Strijbis Foundation (1816199), and two grants from the Dutch Cancer Society (UU2015-7743, RUG-2017-11352).

## Author contributions (CRediT taxonomy)

Conceptualization: CW, JEVI, RZ, PW, ES

Methodology: CW, JEVI, OK, HC, WK, ES

Software: JEVI, LN

Validation: CW, NH, ES

Formal analysis: CW, JEVI, NH, CV, TJ, PD, ES

Investigation: CW, NH, KL, OK, ES

Resources: CW, JEVI, KL, OK, TJ, LN, RZ, PW, ES

Data curation: CW, JEVI, ES

Writing - original draft preparation: CW, ES

Writing - review and editing: All authors

Visualization: CW, JEVI, ES

Supervision: WK, EC, HS, RZ, PW, ES

Project administration: CW, ES

Funding acquisition: HC, WK, RZ, PW

## Supplementary figure legends

**Supplementary figure 1. Tumor specimen and PDO HG-26 prior to rapid drug screening**. A) Macroscopic image of tumor specimen of HG-26 obtained upon recurrence at palliative debulking surgery. Cross-section of the uterus, with exophytic and infiltrating growing tumor, obliterating the uterine cavity. Tumor is perforating deeply into the myometrium. Tumor sample (0.5 cm^3^) was obtained for organoid culture. B-C) Representative brightfield images of PDO HG-26 at day 15.

**Supplementary figure 2. OC PDOs retained genomic features of the original tumor lesion (SNVs)**. Stacked bar chart showing the number of shared (red) and unique (tumor-green, PDO-blue) SNVs between tumor and PDO pairs.

**Supplementary figure 3. OC PDOs retained genomic features of the original tumor lesion (CNAs)**. A) Comparison of genome-wide CNAs in tumor and PDO pairs. B) Genome-wide CNAs in PDO-only samples. Copy-number losses are depicted in blue and gains in red.

**Supplementary figure 4. Correlation of OC PDO drug response with specific measures of clinical response**. (A-E) Drug dose-response curves of OC PDOs for carboplatin and paclitaxel combination treatment dichotomized into clinical response groups. Upper x-axis: carboplatin drug concentrations, lower x-axis: paclitaxel drug concentrations. Dose response curves normalized to positive (navitoclax, ABT-263) and negative controls (DMSO). Non-linear regression analysis: log(inhibitor) vs. normalized response -- Variable slope fit. Red=clinically resistant, blue=clinically sensitive, A) Histopathological tumor response: CRS1=no or minimal response vs CRS2=appreciable response. B) Biochemical response: no normalization (<35 kU/L) of serum CA-125 during primary treatment vs normalization. C) Radiological response: stable disease vs partial response according to RECIST criteria. D) Progression-free survival: <6 months vs >=6 months. E) Overall survival: <17 months vs >=17 months. *Statistically significant difference between sensitive and resistant group according to Wilcoxon signed-rank test corrected for multiple testing (p<0.01).

**Supplementary figure 5. Reproducibility between PDO technical replicates in terms of drug response**. Scatterplot of AUC values for all technical replicates of drug screening data.

**Supplementary figure 6. Likelihood of HR-deficiency based on HR-classifier CHORD**. CHORD-classifier scores for OC PDOs. The dashed line indicates the cut-off for HR-deficiency (<0.5 HR-proficient and >0.5 HR-deficient).

**Supplementary figure 7. Reproducibility between PDO biological replicates in terms of drug response**. A) Scatterplot of IC50-values for all biological replicates (different passage numbers) for 12 drugs and four drug combination treatments. B) Fold-change in IC50-value between the biological replicates. IC50-values were extracted from the drug dose-response curves. A ten-fold change in IC50-value was chosen as an arbitrary cut-off for differential drug response (red dashed line).

**Supplementary figure 8. Drugs that elicit similar drug responses in related OC PDOs**. IC50-values (extracted from dose-response curves) for drugs that elicit similar drug response in all related OC PDOs: carboplatin+adavosertib (A), carboplatin+gemcitabine (B), carboplatin+olaparib (C), carboplatin (D), olaparib niraparib (E), rucaparib (F), alpelisib (G), AZD8055 (H), flavopiridol (I), pictilisib (J), vemurafenib (K). A color code for each patient is shown. The dotted line indicates the steady state (C*ss*) or maximum (C*max*) in vivo plasma concentrations for all single drug treatments (supplementary table 4).

**Supplementary figure 9. Confirmation of *KRAS* mutation status by Sanger sequencing in PDOs MC-3**. Sequencing chromatogram for confirmation of *KRAS* mutation p.G12V in MC-3.1 and −3.2 and p.L19F in MC-3.4.

**Supplementary figure 10. OC PDOs exhibit varying degrees of genome-wide heterogeneity at both the SNV and CNA level** A-H) related OC PDOs with from left to right venn diagrams showing the overlap of all identified SNVs, deletions, and amplifications. In parentheses, the percentage of unique variants in each PDO.

## List of supplementary tables

**Supplementary table 1**. Description of study cohort

**Supplementary table 2**. Clinicopathological data for seven OC PDOs

**Supplementary table 3**. Survival data for HGSOC from the Dutch Cancer Registration

**Supplementary table 4**. The maximum (C*max*) or steady state (C*ss*) *in vivo* plasma concentrations for *in vitro* tested drugs

**Supplementary table 5**. IC50-values per drug for OC PDOs

**Supplementary table 6**. IC50-values for all biological replicates

**Supplementary table 7**. Drug response associated genes from the DGIdb resource

**Supplementary table 8**. SNVs in drug response associated genes in OC PDOs

**Supplementary table 9**. CNAs in drug response associated genes in OC PDOs

